# MOG Antibody Status Shapes Divergent Clinical Profiles And Therapeutic Responses In Chronic Relapsing Inflammatory Optic Neuropathy

**DOI:** 10.64898/2026.04.20.26351249

**Authors:** M. Graure, N. Nierobisch, A. J. De Vere-Tyndall, I. T. Pakeerathan, I. Ayzenberg, J. A. Gernert, J. Havla, M. Ringelstein, O. Aktas, D. Tkachenko, M. W. Hümmert, C. Trebst, N. Cerdá-Fuertes, A. Papadopoulou, K. Giglhuber, R. Wicklein, A. Berthele, M. Weller, V. Kana, P. Roth, M. Herwerth

## Abstract

**Background:** Chronic relapsing inflammatory optic neuropathy (CRION) is a steroid-dependent form of optic neuritis with incompletely understood pathophysiology. The identification of myelin oligodendrocyte glycoprotein antibodies (MOG-IgG) in a substantial patient subset has challenged the diagnostic and therapeutic management. The aim of this study was to investigate clinical profiles and treatment outcomes of patients with CRION, comparing MOG-IgG-positive (MOG+) and seronegative (MOG-) subgroups.

**Methods:** Patients from six European tertiary centers fulfilling diagnostic criteria for CRION were included. All underwent cell-based autoantibody testing. Clinical outcomes (visual acuity, annualized relapse rate), laboratory and imaging findings (MRI, OCT), and treatment responses were retrospectively analyzed.

**Results:** Sixty patients were included (median age 33 years; 70% female); 27 (45%) were MOG+. MOG+ CRION was associated with later onset, higher ARR before treatment (median [IQR] 2 [1-3] vs. 1 [1-2], *p =* 0.023), and a trend toward shorter inter-relapse intervals. Additional distinguishing features included higher frequencies of antinuclear antibody positivity, elevated CSF interleukin-6, and extensive optic neuritis on MRI. Relapse burden correlated with visual acuity decline and retinal thinning.

In MOG+ patients, monoclonal antibody therapy reduced the ARR (n = 21; 2 [1-3] vs. 0 [0-2], p = 0.024), primarily driven by tocilizumab (n = 11; 2 [1-3] vs. 0 [0-1], *p =* 0.023). In MOG-patients, rituximab and azathioprine showed a trend toward ARR reduction.

**Conclusion:** CRION represents a heterogeneous syndrome encompassing distinct subgroups. MOG+ patients demonstrate higher disease activity but respond favorably to tocilizumab. Serological testing is critical for treatment stratification and preventing relapses.

## INTRODUCTION

Chronic relapsing inflammatory optic neuropathy (CRION), a rare, steroid-dependent form of optic neuritis (ON) remains poorly understood. ^1–3^ A key challenge is that, even after excluding alternative diagnoses, CRION is identified only when acute therapy is withdrawn leading to a pathognomonic relapse, rather than at presentation. Steroid tapering in the treatment of ON is not routinely practiced, ^4^ potentially leaving high risk patients vulnerable to preventable relapses. This diagnostic delay poses a significant risk of irreversible visual acuity (VA) worsening and highlights the urgent need for a better characterization and management of this rare disease.

Advances in antibody testing, particularly for aquaporin-4 (AQP4-IgG) and myelin oligodendrocyte glycoprotein (MOG-IgG), have raised debate over whether CRION is a distinct entity, as prior studies found MOG-IgG positivity (MOG+) in 25-90%, and AQP4-IgG seropositivity in 5-25% of patients fulfilling the diagnostic criteria for CRION. ^5–7^

Recommendations for maintenance steroid-sparing therapy remain a considerable challenge in clinical practice and are based on case reports or monocentric studies with no standardized treatment protocols.^8,9^ A spectrum of immunosuppressive agents, monoclonal antibodies, and intravenous immunoglobulins (IVIG) have been proposed as therapeutic options. ^10–13^ However, longitudinal clinical characterization, therapy response and outcome of CRION subtypes remain underexplored.

The aim of this retrospective, longitudinal, multicenter study was to compare clinical profiles and treatment outcomes in patients with the MOG+ versus MOG-CRION phenotypes.

## MATERIALS AND METHODS

### Study design and patient selection

This retrospective, longitudinal, multicenter study included patients >16 years from six tertiary referral centers in Switzerland (n = 1) and Germany (n = 5) who met the diagnostic criteria for CRION.^2^ Medical records and the Neuromyelitis Optica study group database were screened for the keywords “CRION,” “chronic relapsing inflammatory optic neuropathy,” or “relapsing optic neuritis”.

A relapse was defined as an objective worsening in VA persisting for ≥ 24 hours in the absence of an alternative etiology (e.g. infection) and supported by clinical and/or radiological findings. Relapses within 2 months were classified as early relapses. Completeness of records required documentation of VA assessment, at least one brain MRI, MOG-IgG and AQP4-IgG testing, screening for rheumatologic and infectious diseases, and CSF analysis.

Exclusion criteria included demyelinating lesions outside the optic nerves, diagnoses of multiple sclerosis, AQP4-IgG-positive or seronegative neuromyelitis optica spectrum disorder, rheumatologic or infectious disease, as well as incomplete diagnostic workup. Due to the retrospective design, some data were only available for a subset of patients.

For comparison of optical coherence tomography (OCT) outcomes, 30 age- and sex-matched healthy controls without neurological or ophthalmological disease were recruited from a Swiss tertiary referral center.

### Clinical and laboratory data

Except for one individual, all patients underwent at least two independent cell-based assays for MOG-IgG; all patients were screened for AQP4-IgG. Baseline testing was conducted prior to the administration of acute relapse therapies, such as IVIG or corticosteroids. Neutrophil-to-lymphocyte ratio (NLR) was determined within 28 days of ON onset and prior to steroid treatment. If unavailable for the first attack, the earliest NLR data from a subsequent relapse were collected. CSF analysis included cell count and differentiation, albumin quotient, oligoclonal bands, lactate, and CSF/serum glucose ratio. CSF MOG-IgG screening was not routinely performed.

### Magnetic Resonance Imaging

The first available 3 Tesla MRI scans were independently reviewed by two neuroradiologists blinded for the MOG antibody status. Protocols included T1-weighted (± gadolinium), T2-weighted, and FLAIR brain sequences, as well as gadolinium-enhanced and fat-suppressed T2-weighted orbital sequences. Optic nerve localization and extent, perineural involvement, and extra-orbital parenchymal lesions were assessed.

### Visual Acuity

High-contrast visual acuity VA was measured using standardized charts (ETDRS or Snellen). To assess long-term disease burden, only measurements ≥8 weeks after the most recent ON episode were included.

### Optical Coherence Tomography

Retinal imaging was performed with the SPECTRALIS SD-OCT (Heidelberg Engineering) with the automatic real-time function. Peripapillary retinal nerve fibre layer (pRNFL) was assessed with a 3.4 mm ring scan, 12°, centered on the optic nerve head. The common layer of the ganglion cell layer and the inner plexiform layer (GCIPL) was evaluated from macular scans (61 B-scans, scanning angle 30° × 25°) centered on the fovea. Volumes were calculated by the software’s segmentation algorithm and converted into thickness through division by the scanned surface area. Scan quality was verified with OSCAR-IB criteria.^14^

Cross-sectional analyses used the most recent OCT ≥8 weeks after ON onset; longitudinal data were available for a subset of patients.

### Treatment Effectiveness

Therapies were classified as acute (corticosteroids, plasmapheresis) or long-term steroid-sparing. The latter included azathioprine, rituximab, tocilizumab, mycophenolic acid, IVIG, and other agents. For statistical analysis, regimens with ≥5 documented cases were grouped into monoclonal antibodies (rituximab, tocilizumab) and immunosuppressants (azathioprine, mycophenolic acid).

### Statistical analysis

Statistical analyses were performed using R 4.3.2 and GraphPad Prism 10.3.1. Nonparametric tests were applied due to small subgroup sizes and non-normal distributions. Differences were considered statistically significant at *p* < 0.05. Results are presented as medians with interquartile ranges [IQR], or as absolute numbers and percentages of total. Fisher’s exact test was used for categorical variables, Mann-Whitney U for independent samples, and Wilcoxon signed-rank for paired data. Correlations between continuous variables (e.g., VA and number of ON episodes) were assessed by linear regression. The association of multiple predictors with an outcome was assessed using a linear mixed-effects model.

VA outcome analyses were limited to affected eyes. Individual trajectories under steroid-sparing therapy were visualized via swimmer plots; patients with unknown relapse intervals were excluded.

The annualized relapse rate (ARR) for each patient was calculated as number of relapses per year. To avoid possible overestimation of therapy efficacy, patients without ≥12 months of follow-up after therapy initiation were excluded. ARR before and after therapy was compared by mechanism of action (monoclonal antibodies, immunosuppressants), and for individual regimens with ≥5 cases of use respectively.

### Ethical Considerations

Ethical approval was obtained from the Ethics Board of the Canton of Zurich (KEK-ZH-Nr. 2024-00239). All participants provided written informed consent.

## RESULTS

### Study Participants

Sixty patients who met the diagnostic criteria for CRION were included in the study. Of these, 27 individuals (45%) tested positive for MOG-IgG. Among the seronegative patients (*n =* 33/60; 55%), no autoantibodies (MOG-IgG, AQP4-IgG) were detected despite repeated testing.

Both MOG+ and MOG-CRION patients (Table 1) were predominantly female, exhibited a median of 3 ON episodes, and presented with similar low percentage of eyes classified as unaffected by ON. MOG+ CRION individuals were older at onset (median [IQR] 38 years [32-50], vs. 28 years [24.5-39.5] for MOG-, *p =* 0.015; Figure 1A). While the median ARR over the total follow-up period was similar between both subgroups (MOG+: 0.9 [0.5-1.2], MOG-: 0.8 [0.5-1.3]), the median [IQR] ARR in the 12 months before therapy initiation in the MOG+ CRION group was significantly higher compared to the MOG-CRION subgroup (2 [1-3] vs. 1 [1-2], *p =* 0.023; Figure 1B). Early recurrence of ON (inter-ON interval < 2 months) was observed in 39.3% of the total cohort with a trend toward higher occurrence in the MOG+ vs. the MOG-subgroup (14/26 [53.9] vs. 8/30 [26.7]; *p =* 0.055; Figure 1C).

**Figure 1.**
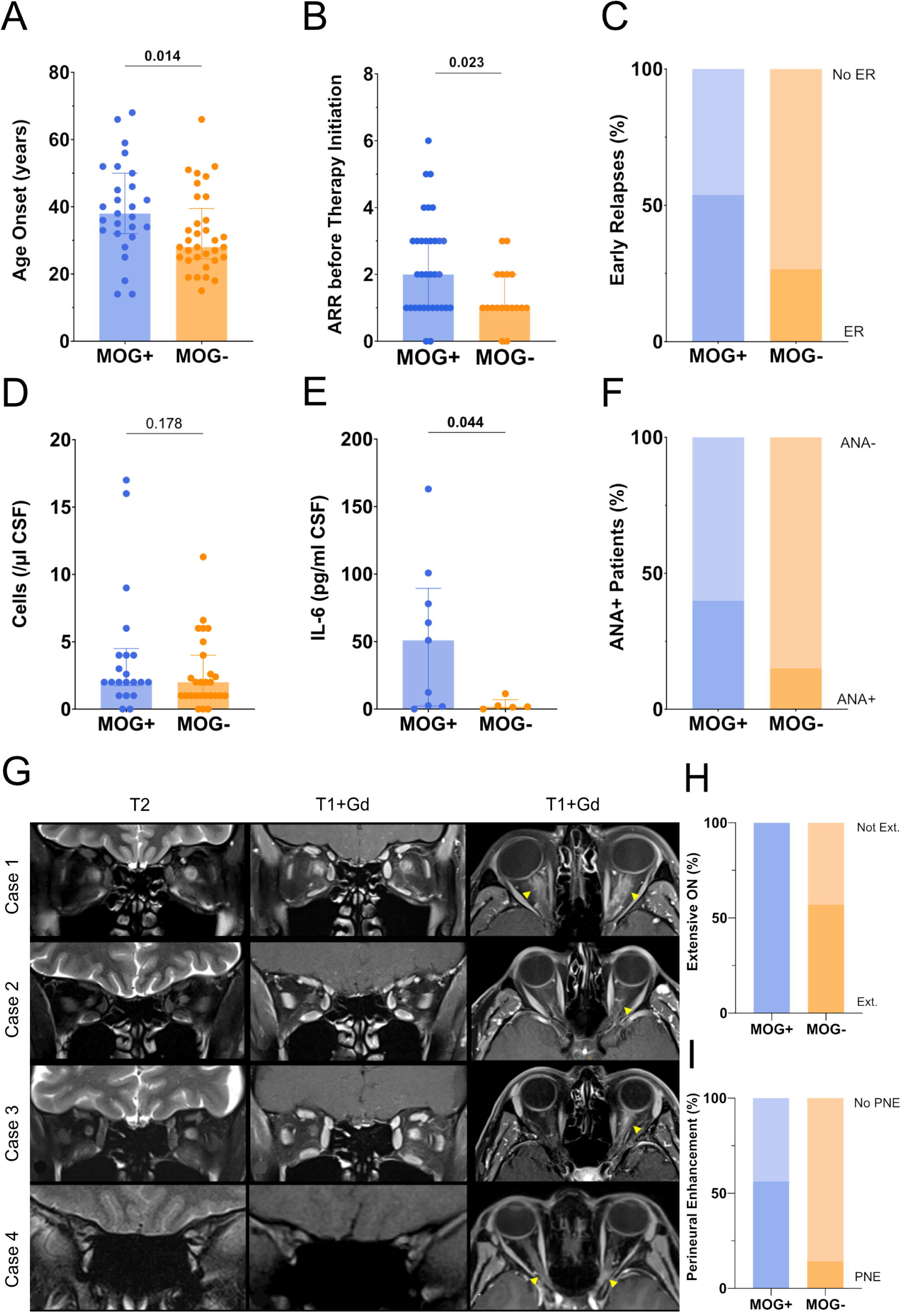
MOG-IgG Status-associated clinical, laboratory, and MRI differences in patients with CRION. (**A**) Bar plot illustrating the median and interquartile range of age at onset (years) and (**B**) the ARR 1 year pre-treatment (Mann-Whitney test). (**C**) Stacked bar plot illustrating percentage of patients with early relapses (Fisher’s exact test). (**D**) Bar plot showing distribution of white blood cell count and (**E**) CSF IL-6 concentrations (Mann-Whitney test). (**F**) Stacked bar plot illustrating percentage of patients positive for antinuclear antibodies (Fisher’s exact test). (***G***) T2-weighted fat-suppressed orbital MRI sequences in the coronal plane (T2, left column), and contrast enhanced T1-weighted fat-suppressed orbital MRI sequences in coronal and axial planes (T1+Gd, middle and right columns). Neural edema appears as hyperintense optic nerve signal alteration on T2-weighted images, whereas neuritic and perineuritic inflammation is reflected as increased contrast enhancement in the T1-weighted images after gadolinium administration (highlighted by yellow arrowheads). Case 1: MOG+ CRION patient with extensive bilateral ON and perineuritis. Case 2: MOG-CRION patient with short subtle left sided ON. Case 3: MOG-CRION patient with extensive fulminant left sided ON with perineuritis. Case 4: MOG-CRION patient with subtle predominantly left sided bilateral optic neuritis. (***H***) Stacked bar plot illustrating percentage of patients demonstrating extensive ON, and (***I***) perineural enhancement by MOG-IgG status at onset (Fisher’s exact test, p = 0.020 and p = 0.089 respectively). Data are presented as median and IQR in A, C, E and F. Each dot represents a single individual. ANA+ = patients who tested positive for antinuclear antibodies; ARR = annualized relapse rate; ER = early relapses; Ext. = extensive; Il-6 =interleukin 6; MOG+= CRION patients who tested positive for MOG-IgG; MOG-= CRION patients who tested negative for MOG-IgG; NLR = neutrophil-to-lymphocyte ratio; ON = optic neuritis; PNE = perineural enhancement.

**Table 1.** Demographic, clinical, laboratory and MRI findings in MOG+ versus MOG-CRION.

| **N (%)** | **Total CRION**<br>**60 (100)** | **MOG+**<br>**27 (45)** | **MOG-**<br>**33 (55)** | **P** |
| --- | --- | --- | --- | --- |
| **Clinical Parameters** |  |  |  |  |
| Sex F, n/N (%) | 42 (70) | 17 (63) | 25 (75.8) | 0.397 |
| Age Onset, yr, median [IQR] | 33 [25.3-44.5] | 38 [32-50] | 28 [24.5-39.5] | **0.015** |
| Total FU, mo, median [IQR] | 50.5 [20.5-97.5] | 56 [13-109] | 45 [25-90.5] | 0.760 |
| ON episodes, median [IQR] | 3 [2-5] | 3 [2-6.3] | 3 [2-4] | 0.264 |
| NON-Eyes, n/N (%) | 13/120 (10.8) | 5/54 (9.3) | 8/66 (12.1) | 0.770 |
| ARR/total FU, median [IQR] | 0.8 [0.5-1.3] | 0.9 [0.5-1.2] | 0.8 [0.5-1.3] | 0.916 |
| ARR 12 months before therapy, median [IQR] (n = 51) | 2 [1-3] | 2 [1-3] | 1 [1-2] | **0.023** |
| Patients with early recurrent ON<sup>a</sup>, n/N (%) | 22/56 (39.3) | 14/26 (53.9) | 8/30 (26.7) | 0.055 |
| VA outcome affected eyes, decimal, median [IQR] (n = 71) | 0.8 [0.4-1] | 0.9 [0.4-1] | 0.8 [0.3-0.9] | 0.358 |
| **Laboratory findings at baseline** |  |  |  |  |
| **Serum** |  |  |  |  |
| Positive ANA, n/N (%) | 15/53 (28.3) | 10/25 (40) | 5/33 (15.2) | **0.040** |
| NLR, median [IQR] (n = 20) | 2.4 [2.1-3.1] | 2.4 [2.2-3.5] | 2.1 [1.9-2.7] | 0.289 |
| **Cerebrospinal fluid** |  |  |  |  |
| Elevated WBCC, n/N (%) | 9/50 (18) | 5/22 (22.7) | 4/28 (14.3) | 0.482 |
| WBCC/μl, median [IQR] (n = 50) | 2 [1-4] | 2 [1.8-4.5] | 2 [1-4] | 0.178 |
| Neutrophil detection, n/N (%) | 3/24 (12.5) | 3/10 (30) | 0/14 (0) | 0.059 |
| Elevated QAlb, n/N (%) | 12/51 (23.5) | 7/22 (31.8) | 5/29 (17.2) | 0.320 |
| Positive OCB, n/N (%) | 9/58 (15.5) | 3/25 (12) | 6/33 (18.2) | 0.718 |
| Elevated Lactate, n/N (%) | 3/35 (8.6) | 2/16 (12.5) | 1/19 (5.3) | 0.582 |
| Low Glucose Index, n/N (%) | 1/33 (3) | 0/14 (0) | 1/19 (5.3) | >0.999 |
| IL-6 pg/ml, median [IQR] (n = 14) | 7 [1.6-67.5] | 50.9 [2.3-89] | 1.7 [0.7-7] | **0.044** |
| **MRI findings at baseline** |  |  |  |  |
| **Dedicated orbital scans N (%)** | 22/23 (95.7%) | 15/16 (93.8%) | 7/7 (100%) | >0.999 |
| Contrast enhancement of optic nerve, n/N (%) | 20/23 (87%) | 14/16 (87.5%) | 6/7 (85.7%) | >0.999 |
| T2 hyperintensity of optic nerve, n/N (%) | 21/23 (91.3%) | 15/16 (93.8%) | 6/7 (85.7%) | 0.526 |
| **ON Localization** |  |  |  |  |
| Orbital, n/N (%) | 22/23 (95.7%) | 16/16 (100%) | 6/7 (85.7%) | 0.304 |
| Canalicular, n/N (%) | 23/23 (100%) | 16/16 (100%) | 7/7 (100%) | >0.999 |
| Chiasmal, n/N (%) | 6/23 (26.1%) | 4/16 (25%) | 2/7 (28.6%) | >0.999 |
| **ON Characteristics** |  |  |  |  |
| Extensive (>1/2), n/N (%) | 20/23 (87%) | 16/16 (100%) | 4/7 (57.1%) | **0.020** |
| Bilateral, n/N (%) | 5/23 (21.7%) | 4/16 (25%) | 1/7 (14.3%) | 1.000 |
| Perineural enhancement, n/N (%) | 10/23 (43.5%) | 9/16 (56.3%) | 1/7 (14.3%) | 0.089 |

<sup>a</sup>Patients with early recurrent ON = patients in which at least 1 interval between relapses was < 2 months. ANA = antinuclear antibodies; ARR = annualized relapse rate; IQR = interquartile range; FU = follow-up; IL-6 = Interleukin 6; mo = months; MOG+ = CRION patients who tested positive for MOG-IgG; MOG- = CRION patients who tested negative for MOG-IgG; n/N = number of cases (n) out of the total number assessed (N); NLR = neutrophil-to-lymphocyte ratio; NON-Eyes = eyes without a history of ON; OCB = oligoclonal bands; ON = optic neuritis episode; QAlb = cerebrospinal fluid/ serum albumin quotient; T2 = T2-weighted fat-suppressed MRI sequence; VA = visual acuity; WBCC = white blood cell count; yr = years.

### Laboratory Findings

A significantly higher proportion of MOG+ CRION individuals showed elevated antinuclear antibody (ANA) titers (10/25; 40%) compared to the MOG-CRION subgroup (5/33; 15.2%; *p* = 0.040; Figure 1F). ANA positivity did not correlate with the number of relapses (*p =* 0.946). Measurement of the NLR within 28 days of relapse onset trended towards higher values in the MOG+ CRION compared to the MOG-CRION subgroup (*n =* 20; median [IQR] 2.4 [2.2-3.5] for MOG+ vs. 2.1 [1.9-2.7] for MOG-; *p* = 0.289).

CSF analysis revealed pleocytosis in 9/50 patients (18%) of the total CRION cohort, with comparable frequencies and a tendency towards higher cell counts in the MOG+ subgroup (Figure 1D). Positive oligoclonal bands were detected in a minority of patients with similar distribution across subgroups. The MOG+ subgroup exhibited significantly higher CSF IL-6 levels than MOG-CRION patients (*n =* 14; median [IQR] for MOG+ 50.9 [2.3-89] vs. 1.7 [0.7-7] in MOG-; *p =* 0.044; Figure 1E). Neutrophils were observed in the CSF of MOG+ individuals but were absent in the MOG-subgroup (3/10 [30%] for MOG+ vs. 0/14 [0%] for MOG-; *p =* 0.059).

### Radiological Findings

MRI data were available for 23 patients. Dedicated orbital MRI scans (T1-weighted, gadolinium enhanced fat-suppressed T1-weighted, fat-suppressed T2-weighted and FLAIR) were available for most patients (95.7%). Extensive optic nerve involvement, defined as lesions affecting more than half of the nerve length, was significantly more common in MOG+ CRION individuals compared to the MOG-subgroup (16/16 [100%] vs 4/7 [57.1%]; *p =* 0.020, Table 1; Figure 1H). MOG+ CRION cases exhibited a higher tendency for perineural contrast enhancement compared to MOG-cases, though the difference did not reach statistical significance (9/16 [56.3%] vs. 1/7 [14.3%]; *p =* 0.089; Figure 1I).

There were no differences in localization patterns between the MOG+ and MOG-CRION subgroups (Table 1). All patients (100%) exhibited involvement of the canalicular segment, while the orbital segment was affected in most cases (95.7%). Affection of the optic chiasm was observed in 6 patients (26.1%).

### Visual Outcome

Analysis of VA of affected eyes (*n =* 71) in individuals with CRION revealed that a higher number of ON episodes correlated with poorer VA outcomes (B = −0.036, 95% confidence intervals [CI] −0.065 to −0.006, *p =* 0.018), while worsening was accentuated in the MOG+ subgroup (Figure 2). Early relapse recurrence or higher IL-6 levels were not predictive of poorer VA outcome (*p =* 0.679 and *p =* 0.090, respectively), though the latter showed a potential trend.

**Figure 2.**
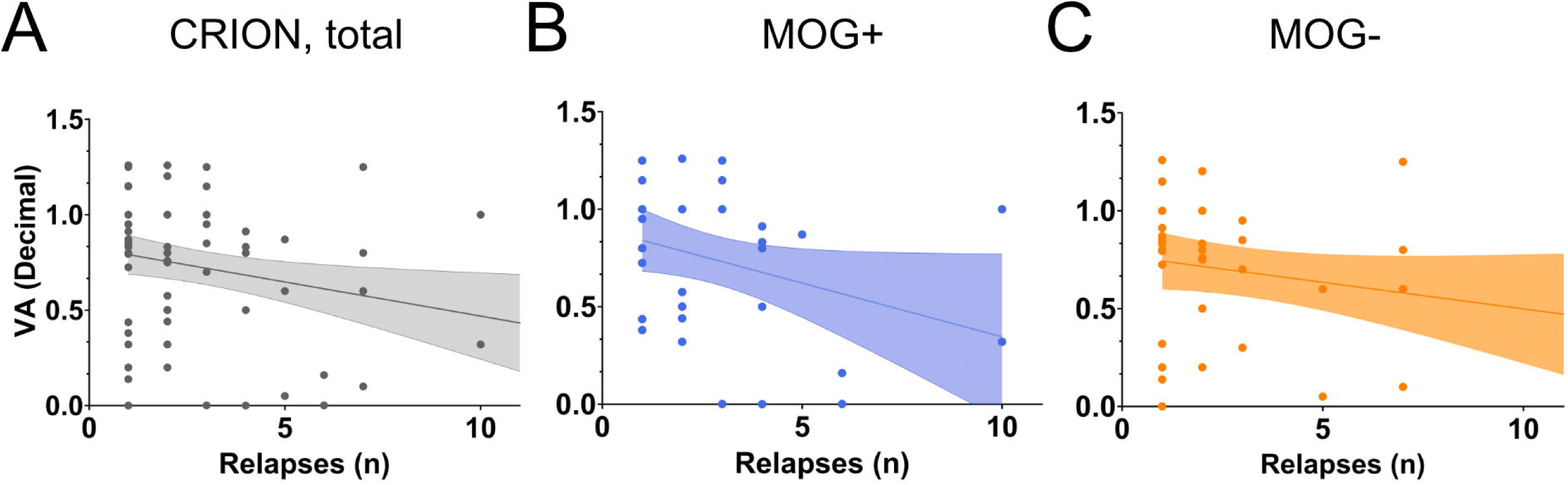
Visual acuity outcome of affected eyes in patients with CRION. (**A-C**) Scatterplots with linear regression lines and 95% CI (light-shaded bands) illustrating the relationship between the visual acuity outcome (decimal) and the number of relapses (**A**) in the total cohort, (**B**) in the MOG+ subgroup, and (**C**) in the MOG-subgroup. Simple linear regression, *p =* 0.018, (A), *p =* 0.055 (B), *p =* 0.126 (C); each dot represents an affected eye. MOG+= CRION patients who tested positive for MOG-IgG; MOG-= CRION patients who tested negative for MOG-IgG; *n =* number; VA = visual acuity.

### Optical Coherence Tomography Findings

In both CRION subgroups, pRNFL and GCIPL thicknesses were significantly lower in eyes with ON compared to age- and sex-matched healthy controls. The pRNFL thickness showed progressive decline with increasing number of ON across groups (pRNFL in MOG+: healthy controls vs. 1-ON vs. ≥2 ON, median [IQR] 100 [98-106] vs. 76.5 [60.8-85.5] vs. 55 [39-72], p < 0.001 respectively; pRNFL MOG-: healthy controls vs. 1-ON vs. ≥2 ON, median [IQR] 102 [94-110] vs. 83 [67.5-99.8] vs. 84.5 [44.3-105.8], *p =* 0.004 and 0.005, respectively). A decline was evident for GCIPL thickness (GCPIL MOG+: healthy controls vs. 1-ON vs. ≥2 ON, median [IQR] 86.3 [83.5-91.9] vs. 75 [54.5-83.5] vs. 62.2 [46-77.4], *p =* 0.001 for both comparisons; GCIPL MOG-: healthy controls vs. 1-ON vs. ≥2 ON, median [IQR] 88.4 [82.4-93] vs. 50.9 [28.3-69.3] vs. 67.9 [48.1-84.9], p < 0.001 and *p =* 0.010 respectively; Figure 3A-B and D-E).

**Figure 3.**
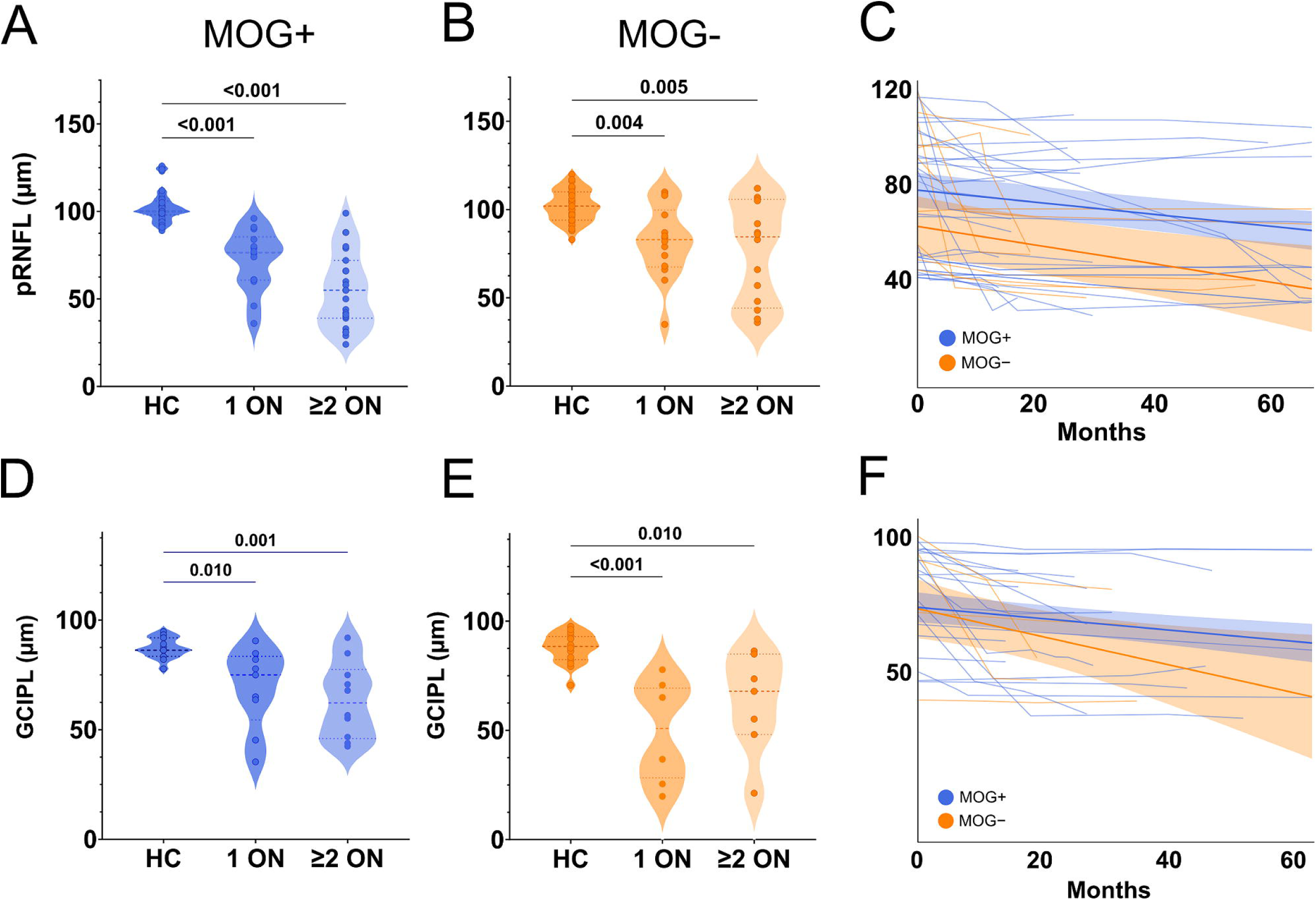
Temporal changes in OCT-based pRNFL and GCIPL thickness across CRION subgroups. (**A-B**) Violin plots illustrating the median, IQR, and range of pRNFL and (**D-E**) GCIPL thickness in µm in healthy controls compared to MOG+ (**A; D**) and MOG-(**B; E**) patients with 1 ON and ≥2 ON respectively (Kruskal-Wallis test, followed post-hoc by Dunn’s test). Each dot represents a single eye. (**C**) Longitudinal pRNFL and (**F**) GCIPL thickness trajectories from MOG+ patients (blue) and MOG-CRION patients (orange) predicted by the mixed linear effect model (thick lines = median; light-shaded bands = 95% CI). Thin lines show individual eye-based trajectories, plotted over the maximum follow-up duration. GCIPL = ganglion cell and inner plexiform thickness; HC = healthy controls; MOG+ = CRION patients who tested positive for MOG-IgG; MOG-= CRION patients who tested negative for MOG-IgG; pRNFL = peripapillary retinal nerve fiber layer.

In patients with longitudinal data (23/60, *n =* 45 eyes), both pRNFL and GCIPL layers showed a progressive thinning in the MOG+ and MOG-subgroups (Figure 3C, F). The high ARR, with a median [IQR] of 0.8 [0.5-1.3] ON attacks per year during the follow-up period, precluded a longitudinal analysis independent of ON episodes. Thicknesses declined for both pRNFL (estimate, B = −0.252, standard error, SE = 0.054, p[<[0.001) and GCIPL (B = −0.178, SE = 0.050, p < 0.001) in MOG+ CRION patients over time, as did pRNFL (B = −0.389, SE = 0.135, p[= 0.005) and GCIPL (B = −0.439, SE = 0.180, *p =* 0.018) in MOG-CRION patients. There was no significant difference in thinning rates between the groups (pRNFL: *p =* 0.347, GCIPL: *p =* 0.167).

### Treatment Choice and Efficacy

All patients received corticosteroids during acute ON episodes and subsequently demonstrated improvement in VA, as required by the inclusion criteria (Table 2). The median number of 3 corticosteroid courses per patient, and the proportion undergoing plasmapheresis, e.g. in severe cases or due to recurrent VA decline post-steroid-tapering, were comparable between subgroups (8/27 [29.6%] for MOG+ vs. 11/33 [33.3%] for MOG-; Table 2).

**Table 2.**
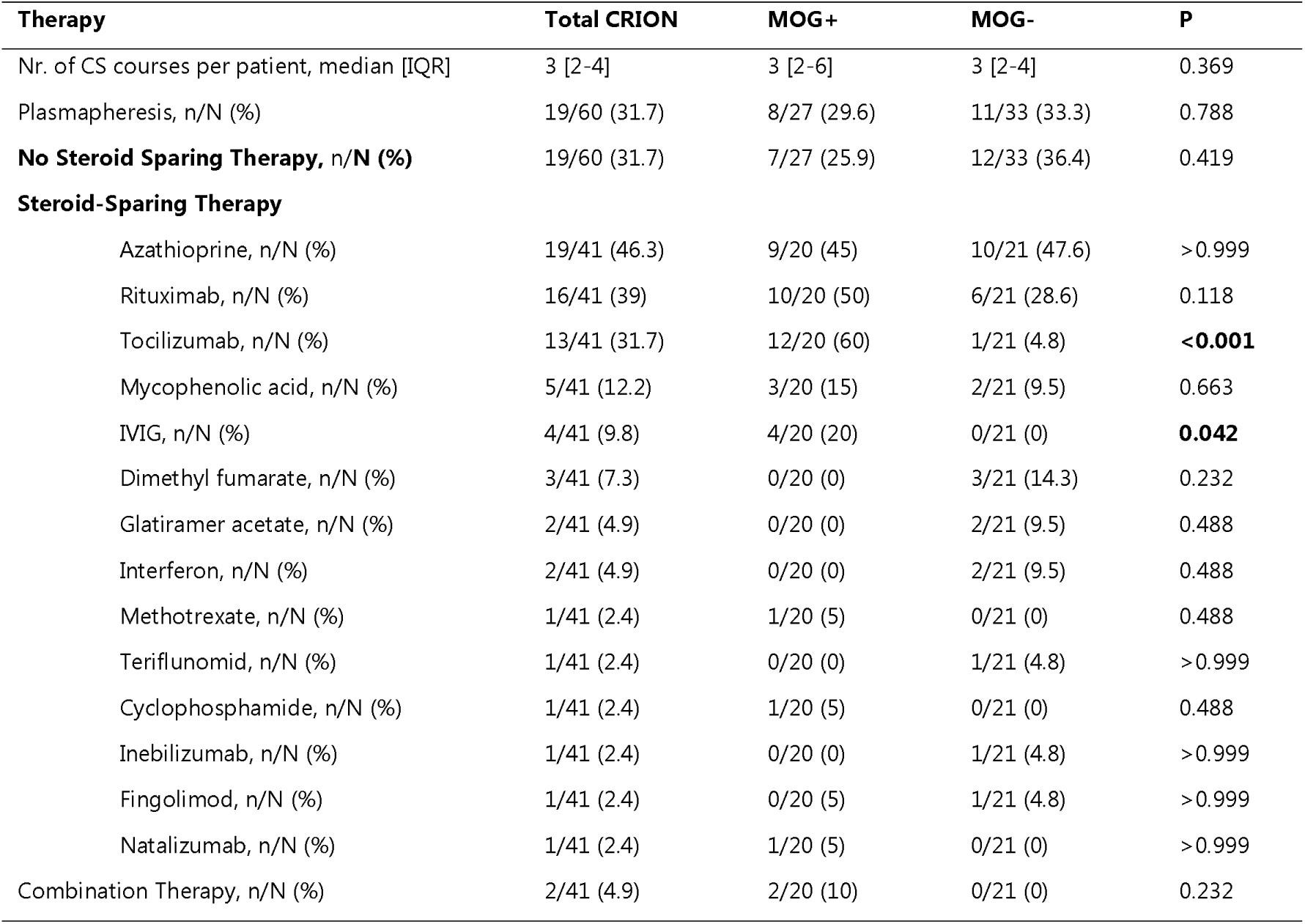
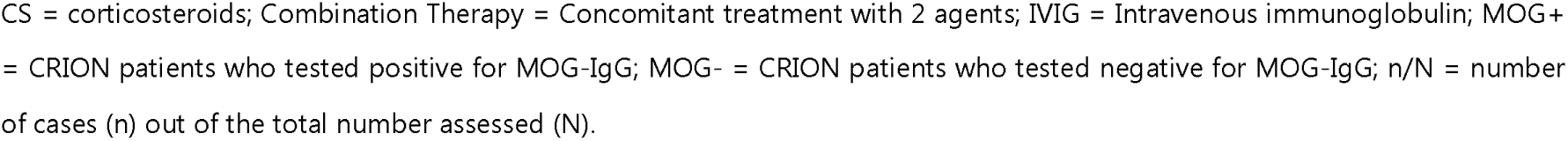
Acute and maintenance therapy of patients with CRION.

| **Therapy** | **Total CRION** | **MOG+** | **MOG-** | **P** |
| --- | --- | --- | --- | --- |
| Nr. of CS courses per patient, median [IQR] | 3 [2-4] | 3 [2-6] | 3 [2-4] | 0.369 |
| Plasmapheresis, n/N (%) | 19/60 (31.7) | 8/27 (29.6) | 11/33 (33.3) | 0.788 |
| **No Steroid Sparing Therapy, n/N (%)** | 19/60 (31.7) | 7/27 (25.9) | 12/33 (36.4) | 0.419 |
| **Steroid-Sparing Therapy** |  |  |  |  |
| Azathioprine, n/N (%) | 19/41 (46.3) | 9/20 (45) | 10/21 (47.6) | >0.999 |
| Rituximab, n/N (%) | 16/41 (39) | 10/20 (50) | 6/21 (28.6) | 0.118 |
| Tocilizumab, n/N (%) | 13/41 (31.7) | 12/20 (60) | 1/21 (4.8) | **<0.001** |
| Mycophenolic acid, n/N (%) | 5/41 (12.2) | 3/20 (15) | 2/21 (9.5) | 0.663 |
| IVIg, n/N (%) | 4/41 (9.8) | 4/20 (20) | 0/21 (0) | **0.042** |
| Dimethyl fumarate, n/N (%) | 3/41 (7.3) | 0/20 (0) | 3/21 (14.3) | 0.232 |
| Glatiramer acetate, n/N (%) | 2/41 (4.9) | 0/20 (0) | 2/21 (9.5) | 0.488 |
| Interferon, n/N (%) | 2/41 (4.9) | 0/20 (0) | 2/21 (9.5) | 0.488 |
| Methotrexate, n/N (%) | 1/41 (2.4) | 1/20 (5) | 0/21 (0) | 0.488 |
| Teriflunomid, n/N (%) | 1/41 (2.4) | 0/20 (0) | 1/21 (4.8) | >0.999 |
| Cyclophosphamide, n/N (%) | 1/41 (2.4) | 1/20 (5) | 0/21 (0) | 0.488 |
| Inebilizumab, n/N (%) | 1/41 (2.4) | 0/20 (0) | 1/21 (4.8) | >0.999 |
| Fingolimod, n/N (%) | 1/41 (2.4) | 0/20 (5) | 1/21 (4.8) | >0.999 |
| Natalizumab, n/N (%) | 1/41 (2.4) | 1/20 (5) | 0/21 (0) | >0.999 |
| Combination Therapy, n/N (%) | 2/41 (4.9) | 2/20 (10) | 0/21 (0) | 0.232 |

CS = corticosteroids; Combination Therapy = Concomitant treatment with 2 agents; IVIG = Intravenous immunoglobulin; MOG+ = CRION patients who tested positive for MOG-IgG; MOG- = CRION patients who tested negative for MOG-IgG; n/N = number of cases (n) out of the total number assessed (N).

19/60 CRION patients (31.7%) did not receive any long-term steroid-sparing therapy (7/27 [25.9] for MOG+ vs. 12/33 [36.4] for MOG-; *p =* 0.419). Adjustment of treatment strategies was required more often in MOG+ CRION cases, with individuals receiving a median [IQR] of 1.5 [1-2.3] therapies vs. 1 [1-1] in MOG-(*p =* 0.041; Figure 4). Azathioprine (19/41 [46.3%]) and rituximab (16/41; [39%]) were the most commonly used agents across both CRION subgroups (Table 2). Tocilizumab was preferentially initiated in MOG+ CRION patients, while IVIG was administered exclusively to individuals in the MOG+ subgroup. Mycophenolic acid was prescribed in (5/41 [12.2%)] of the overall cohort, with similar distribution between subgroups. Dimethyl fumarate was used exclusively in (3/21 [14.3%]) of MOG-CRION patients. Other therapies were used infrequently across the cohort (Table 2). Concomitant treatment with 2 steroid-sparing agents was administered exclusively within the MOG+ CRION subgroup (2/20; [10%]). One patient received a combination of rituximab and mycophenolic acid. Another patient was initially treated with methotrexate with concomitant tocilizumab, which was later discontinued due to relapse; subsequent management included a regimen of methotrexate, IVIG, and low-dose corticosteroids. Individual longitudinal disease courses and maintenance immunotherapies for both MOG+ and MOG-CRION subgroups are illustrated in Figure 4.

**Figure 4.**
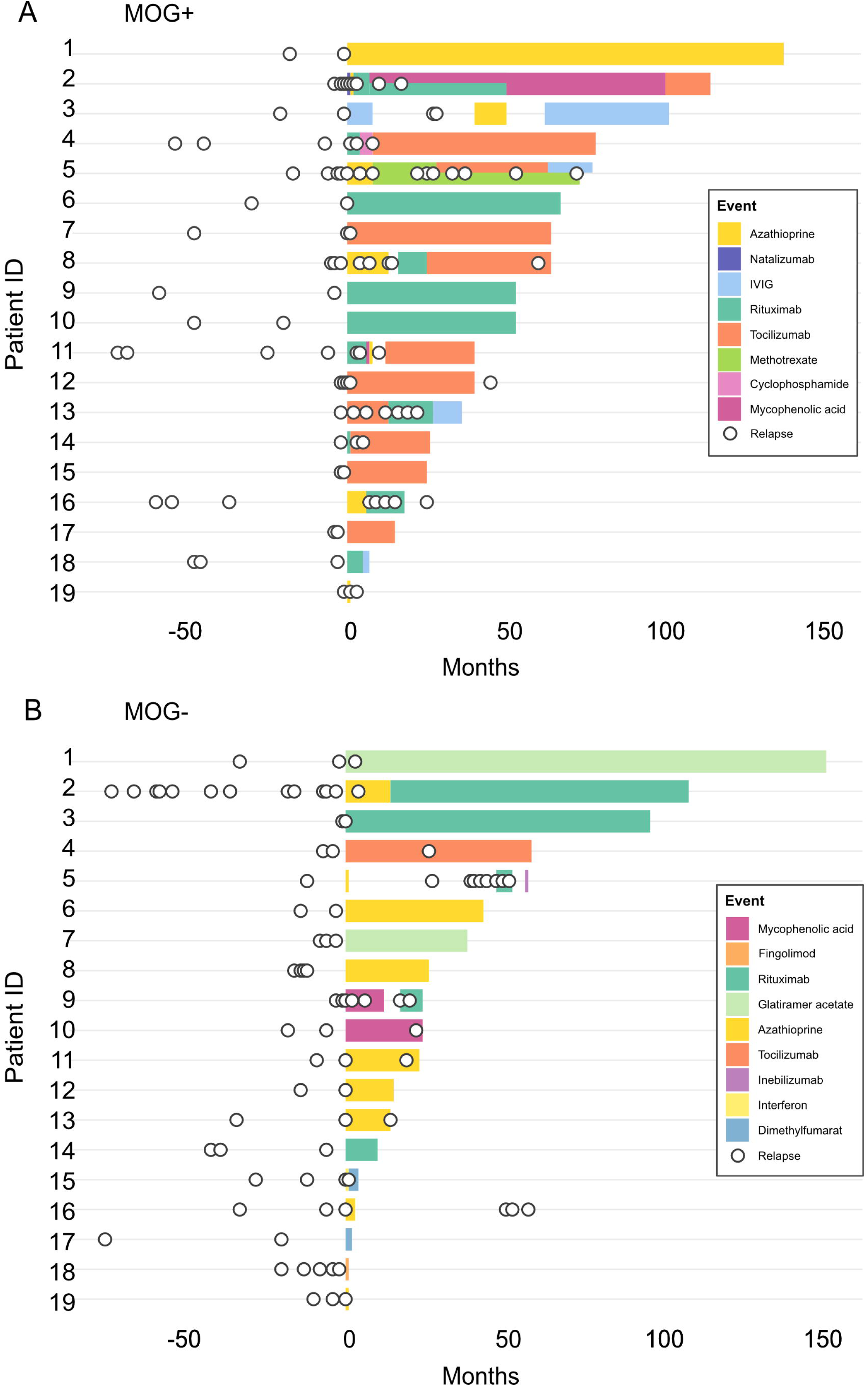
Longitudinal disease courses and individual maintenance therapies across CRION subgroups. (**A; B**) Swimmer plot illustrating individual disease trajectories in MOG+ and MOG-individuals. Attacks are indicated as circles. IVIG = IV immunoglobulin; MOG+ = CRION patients who tested positive for MOG-IgG; MOG-= CRION patients who tested negative for MOG-IgG.

In 28/51 cases (54.9%) there was at least one documented relapse within 12 months after initiation of a new steroid-sparing therapy. ARR for the entire cohort declined significantly following initiation of maintenance therapy, regardless of treatment type (median [IQR] 2 [1-3] pre-treatment vs. 1 [0-2] post-treatment; *p =* 0.003). Analysis of treatment strategies yielded a significant association between initiation of monoclonal antibodies (encompassing rituximab and tocilizumab) in MOG+ patients and ARR reduction (*n =* 21; median [IQR] before vs. after: 2 [1-3] vs. 0 [0-2], *p =* 0.024). In patients receiving immunosuppressive therapy (*n =* 10), the ARR remained unchanged (median [IQR] before 2 [0-3] vs. after 2 [0-3]; *p =* >0.999; Figure 5A).

**Figure 5.**
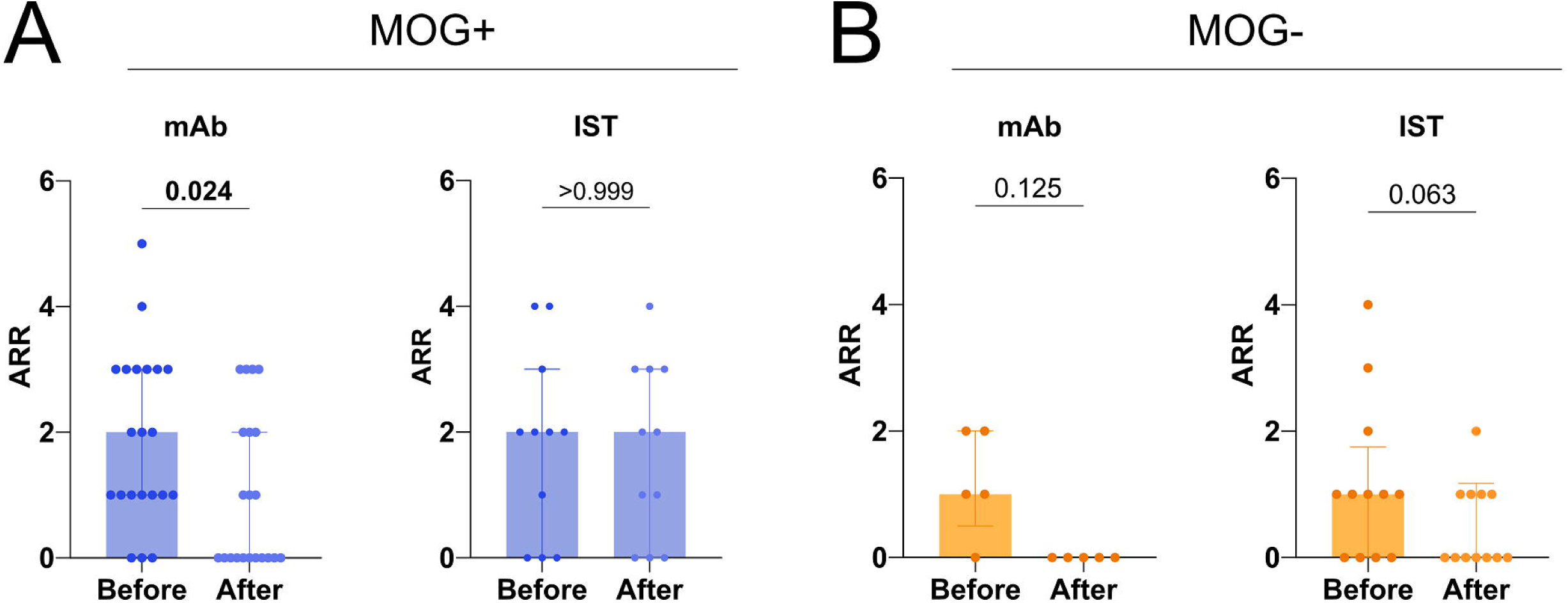
Annualized relapse rate before and after maintenance therapy initiation. (**A; B**) Box plots illustrating median and IQR of the ARR in the MOG+ subgroup (left) and MOG-subgroup (right), comparing the 12 months before and after initiation of the four most used treatment strategies: immunosuppressive therapies (azathioprine, mycophenolic acid) and monoclonal antibodies (rituximab, tocilizumab); Wilcoxon matched pairs signed rank test. Data are presented as median and IQR. Each dot represents a single individual. ARR = annualized relapse rate; IST = immunosuppressive therapies; mAb = monoclonal antibodies; MOG+ = CRION patients positive for anti-myelin oligodendrocyte glycoprotein antibody; MOG-= CRION patients negative for anti-myelin oligodendrocyte glycoprotein antibody.

In contrast, immunosuppressive therapy in MOG-CRION subgroup was associated with a reduction in ARR (*n =* 12; median [IQR] before vs. after 1 [0-2] vs. 0 [0-1], *p =* 0.063). Monoclonal antibody therapy was used less frequently (*n =* 5), but showed a comparable trend toward ARR reduction (median [IQR] 1 [0.5-2] before vs. after 0 [0-0], *p =* 0.125; Figure 5B).

Subgroup analysis revealed that the positive response to monoclonal antibodies in MOG+ CRION was driven by the effect of tocilizumab (*n =* 11; median [IQR] before: 2 [1-3] vs. after: 0 [0-1], *p =* 0.023; Supplementary Figure 1C). In contrast, neither rituximab (*n =* 10) nor azathioprine (*n =* 8) or mycophenolic acid (*n =* 3) was associated with a reduction of ARR in the MOG+ CRION subgroup (median [IQR] before vs. after rituximab: 1 [1-3.3] vs. 1.5 [0-3], *p =* 0.617; before vs. after azathioprine: 2 [0.3-3.8] vs. 2 [0.3-3], p > 0.999; before vs. after mycophenolic acid: 2 [0-2] vs. 1 [0-3], p > 0.999; Supplementary Figure 1A-B,D). In the MOG-subgroup, individuals receiving either rituximab (*n =* 4) or azathioprine (*n =* 9) demonstrated a favorable trend (median [IQR] ARR before vs. after rituximab: 1 [0.3-1.8] vs. 0 [0-0], *p =* 0.250; before vs. after azathioprine: 1 [0-1.5] vs. 0 [0-1], *p =* 0.250; Supplementary Figure 1A-B). The limited use of mycophenolic acid (*n =* 2) and tocilizumab (*n =* 1) precluded further analysis. Treatment selection in the patient receiving IL-6 blocking therapy was guided by evidence of elevated CSF IL-6 levels at disease onset (11.4 pg/µl; the 95th percentile in control subjects is 7.0 pg/ml).

## DISCUSSION

Due to its rarity and the lack of systematic clinical data, CRION remains an underexplored condition, posing substantial diagnostic and therapeutic challenges. In this multicenter retrospective longitudinal study, we compared patients with MOG-IgG-seropositive with MOG-IgG-seronegative CRION and found differences both in disease characteristics and treatment responses between the two groups.

MOG+ CRION patients demonstrated higher disease activity, characterized by elevated relapse rates 12 months prior to treatment initiation and a tendency toward shorter intervals between relapses compared to the MOG-subgroup. Notably, the overall ARR in the CRION cohort (median ARR 0.8, IQR [0.5-1.3]), independent of antibody status, appeared higher compared to the ARR range typically reported for multiple sclerosis (ARR 0.05-0.40),^15,16^ neuromyelitis optica spectrum disorder (ARR 0.1-0.50),^17,18^ and MOGAD in general (ARR 0.05-0.35).^18,19^ This emphasizes the urgent need for early effective treatment approaches in CRION to prevent irreversible visual impairment.

Recurrent episodes of ON lead to cumulative damage to the optic nerve, constituting a key risk factor for poor VA outcomes.^20–22^ Despite prompt treatment and favourable steroid responsiveness, visual acuity declined with increasing numbers of ON episodes across subgroups, reinforcing the importance of relapse prevention as a central therapeutic goal in the management of CRION. Additionally, OCT revealed significant retinal thinning, with progressive pRNFL and GCIPL atrophy in both MOG+ and MOG-CRION patients over time, with no significant intergroup differences. These retinal changes are most likely attributable to sustained relapse activity (median 0.8 ON attacks/year) during the follow-up period, which prevented meaningful assessment of relapse-independent structural progression.

Although some individuals suffered severe and irreversible vision loss, overall VA outcomes were more favourable than previously reported.^2^ This disparity may reflect diagnostic advances enabling earlier identification of CRION, as well as autoantibody-based reclassification, excluding AQP4-IgG positive NMOSD cases typically associated with worse VA outcomes. For example, one study reported an increase in AQP4-IgG seropositivity from 5% to 22% following reanalysis of serum samples from a previously characterized CRION cohort.^7^ The introduction of steroid tapering protocols in atypical ON and the increasing use of monoclonal antibodies, particularly in MOG+ CRION individuals, may further contribute to better long-term outcomes. Finally, our analysis included only assessments performed >8 weeks after symptom onset. Differences in follow-up duration, clinical evaluation, and data reporting across previous studies may account for the observed discrepancies.

Further distinguishing features of MOG+ CRION cases included older age at onset, more extensive optic neuritis involvement, and a trend toward more frequent perineural involvement. Laboratory analysis revealed that markers of systemic autoimmunity and inflammation were more frequently detected in the MOG+ CRION subgroup compared to MOG-CRION patients. This included a higher frequency of antinuclear antibody positivity (40%), exceeding that observed in MOG-patients or previously reported values in MOGAD cohorts with mixed phenotypes (7-24%). ^23–27^ In CSF, neutrophils were observed exclusively in MOG+ individuals and CSF IL-6 levels were significantly elevated compared to MOG-patients. These findings collectively suggest increased disease activity and IL-6 pathway involvement in the MOG+ CRION phenotype, consistent with previous reports on MOGAD^23,24,28,29^, and may guide treatment strategies for this subgroup.

In clinical practice, treatment strategies for MOGAD, seropositive and seronegative NMOSD, and related disorders commonly include both immunosuppressive agents and monoclonal antibodies.^30–34^ Notably, in the MOG+ CRION subgroup, the use of monoclonal antibodies was associated with a significant reduction in ARR, in contrast to the limited response observed with conventional immunosuppressants. Subgroup analysis indicated that the positive effect was mainly driven by tocilizumab, aligning with prior reports for different MOGAD phenotypes.^35,36^ Neither rituximab nor azathioprine significantly reduced the ARR in MOG+ CRION, supporting emerging evidence that rituximab may be less effective in MOGAD.^37,38^

The MOG-CRION subgroup demonstrated a trend toward reduced relapse frequency with both immunosuppressive therapies and monoclonal antibodies, consistent with the limited evidence available from prior studies.^10,11^ Subgroup analysis distinguished both azathioprine and rituximab as potential therapeutic options, however, small sample sizes limit the strength of these conclusions. The restricted use of tocilizumab in MOG-CRION patients precluded assessment of effectiveness.

The initial description of CRION^1^, along with subsequent studies, proposed that CRION is a distinct disease entity.^2,39^ However, after the discovery of MOG-IgG, others have suggested that CRION may represent a manifestation of MOGAD spectrum, based on high MOG-IgG seropositivity rates in two independent cohorts.^5,6^ In our longitudinal cohort, only 45% of patients were ultimately found to be MOG+. This is a lower percentage compared to some previously published data^5^, despite repetitive testing using cell-based assays. Although early relapse has been proposed as a clinical predictor of MOG-IgG seropositivity^5^, approximately 27% of MOG-CRION patients also experienced early relapses, further highlighting the heterogeneity of CRION and the potential involvement of as yet unidentified immune mechanisms. Therefore, our findings suggest the conceptualization of CRION as a potentially distinct clinical syndrome comprising immunologically diverse subgroups with variable treatment responses.

This study has several limitations inherent to its retrospective design, including variable follow-up durations and non-standardized treatment decisions, which were influenced by the treating physicians’ clinical judgment, local drug availability, and cost considerations. Steroid tapering protocols were not standardized across institutions, and different therapy durations may have affected inter-relapse intervals. Additionally, some patients did not receive consistent tapering after every ON, possibly driving VA worsening. There was no systematic assessment of MOG-IgG in the CSF. Considering that CSF-restricted positivity has been described in approximately 0.1-10% of MOGAD cases^40^, a small number of MOG-CRION individuals may represent undetected MOGAD cases. The small size of treatment subgroups limits conclusions on therapy effectiveness in general.

Despite these limitations, this study offers valuable insights into clinical profiles and treatment responses of MOG+ compared to MOG-CRION patients, suggesting that these subgroups may benefit from distinct treatment approaches. These findings support the diagnosis of CRION as a clinical syndrome, likely involving different autoimmune pathways and possibly hitherto undiscovered autoantibody targets. Further research in larger cohorts on the clinical, paraclinical, immunological, and therapeutic response profiles of CRION subgroups is essential to guide personalized treatment strategies and prevent vision loss.

## Author contribution

MG and MH were responsible for the concept and study design. NN, ADVT, ITP, JAG, JH, DT, NCF, KG, and RW contributed to data collection. MG, MH and VK were involved in data analysis. MG and MH drafted the manuscript and figures with the input from all the authors. NN, ADVT, ITP, JAG, JH, DT, NCF, KG, RW, IA, MR, MWH, CT, AP, AB, OA, MW, VK, and PR contributed to writing-review and editing. All authors discussed results and contributed to revision of the manuscript.

## Data Availability

All data produced in the present study are available upon reasonable request to the authors.

## Acknowledgements

We thank D. Agostino and M. Weise for the technical support and OCT image acquisition. MH was supported by the Swiss National Science Foundation (PZ00’3_216616/1), by the Olga-Mayenfisch-Foundation (2024) and by the Betty and David Koetser Foundation. MG was supported by the Betty and David Koetser Foundation. DT was supported by PRACTIS Clinician Scientist Program, funded by Hannover Medical School and DFG (DFG ME 3696/3).

