## Supplementary Figure 1 for "MOG Antibody Status Shapes Divergent Clinical Profiles And Therapeutic Responses In Chronic Relapsing Inflammatory Optic Neuropathy"

### SUPPLEMENTARY MATERIAL

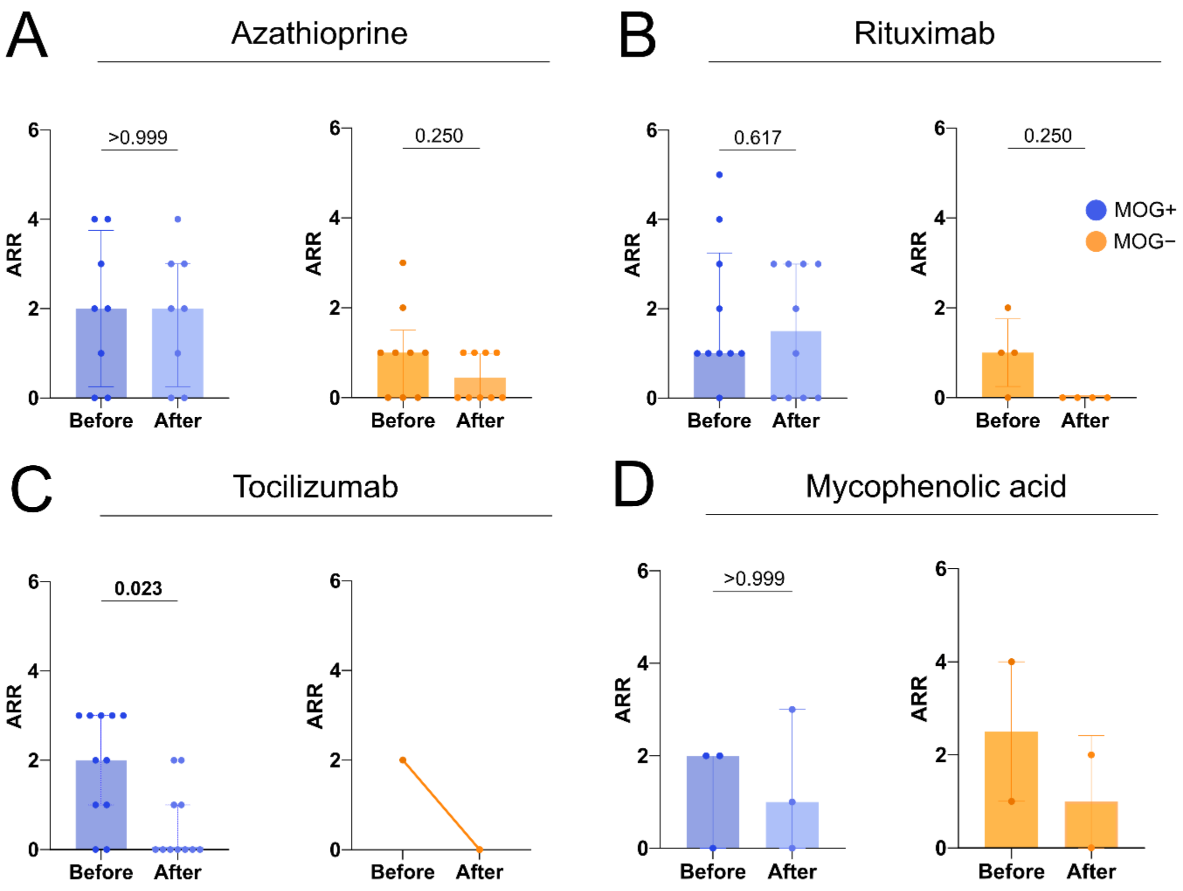


**Supplementary Figure 1 Annualized relapse rate before and after maintenance therapy initiation across CRION subgroups.** (**A-D**) Box plots illustrating the median and IQR of ARR in MOG+ subgroup (left) and MOG- subgroup (right) 12 months before and after initiation of four main therapeutics: (**A**) azathioprine, (**B**) rituximab, (**C**) tocilizumab, and (**D**) mycophenolic acid. Wilcoxon matched pairs signed rank test. Data are presented as median and IQR. Each dot represents a single individual. ARR = annualized relapse rate; MOG+ = CRION patients who tested positive for anti-myelin oligodendrocyte glycoprotein antibody; MOG- = CRION patients who tested negative for anti-myelin oligodendrocyte glycoprotein antibody.
